# Prediction of Imminent Sudden Cardiac Arrest Using a Combination of Warning Symptoms and Clinical Features

**DOI:** 10.1101/2025.03.05.25323376

**Authors:** Kyndaron Reinier, Harpriya Chugh, Arayik Sargsyan, Audrey Uy-Evanado, Kotoka Nakamura, Elizabeth Heckard, Marco Mathias, Tristan Grogan, David Elashoff, Angelo Salvucci, Jonathan Jui, Sumeet S. Chugh

## Abstract

**Introduction:** At least 50% of individuals who suffer sudden cardiac arrest (SCA) experience warning symptoms before their SCA. We have previously reported chest pain and dyspnea as the most common and potentially predictive symptoms. Combining with clinical features could improve sensitivity and specificity for prediction of imminent SCA (ISCA).

**Hypothesis:** A combination of warning symptoms and clinical profiles can predict ISCA.

**Methods:** From two community-based studies of SCA in Oregon and California, we conducted a case-control study. Cases (n=364) were survivors of SCA who had experienced warning symptoms, and control subjects (n=313) were individuals who notified emergency medical services (EMS) for similar symptoms but did not have SCA. Symptom data were obtained from interviews with study subjects and from EMS pre-hospital care records. We constructed classification and regression tree (CART) models for major symptom categories to identify clinical predictors of ISCA. We used the area under the receiver operating characteristic curve (AUC) and 5-fold cross-validation to assess model performance and stability.

**Results:** Heart failure (HF) and/or coronary artery disease (CAD) were predictors of ISCA and displayed important sex differences. For example, among individuals presenting with only chest pain, male sex, particularly males with HF, was an important predictor of ISCA (AUC = 0.813). Among individuals with only dyspnea, CAD and HF were important predictors (AUC = 0.745) with no sex differences identified. The 5-fold cross-validation produced consistent results.

**Conclusions:** Combinations of warning symptoms and clinical features distinguished individuals with SCA from individuals without SCA with good accuracy (AUCs 0.728 – 0.813).

## INTRODUCTION

Out-of-hospital sudden cardiac arrest (SCA), an abrupt cessation of cardiac function causing immediate loss of circulation, breathing, and consciousness,^1^ has a fatality rate >90% and is responsible for more years of life lost than any single cancer in the United States.^2^ Approximately half of SCA patients experience symptoms in the hours before their cardiac arrest, including chest pain, dyspnea, lightheadedness, palpitations, syncope, weakness/fatigue, and others.^3–9^ Ideally, these early warning symptoms could be used to identify individuals at imminent risk of SCA for monitoring and potential early treatment, thus reducing mortality from SCA. In the context of imminent SCA (ISCA), defined here as SCA occurring within hours or days, warning symptoms could be identified early using digital technology^10^ for triage, early intervention and near-term prevention of death from SCA.^11^

In an earlier publication, we reported that among patients with symptoms, those who called 911 before their collapse from SCA (~20% of such patients) had >5-fold higher survival compared to individuals with symptoms who did not.^8^ Subsequently, we conducted a case-control analysis to evaluate the utility of specific symptoms, noted by paramedics in the emergency medical services (EMS) pre-hospital care report, for prediction of ISCA in two established and ongoing community-based studies of out-of-hospital SCA.^12^ In that study, 411 (50%) of 823 witnessed SCA cases had at least one warning symptom on day of the SCA. SCA cases were more likely than individuals in the control group to have dyspnea (40.9% vs. 22.4%), chest pain (33.1% vs. 25.3%), diaphoresis (12.2% vs. 7.7%), and seizure-like activity (10.5% vs. 6.6%) (p<0.01),^12^ though findings differed significantly by sex, with only dyspnea more likely among women. Our findings were consistent in an external replication population. Nonetheless, no specific symptom was sensitive or specific enough to use alone for prediction of ISCA.

We hypothesized that combining symptoms with patient clinical history could improve prediction of ISCA. Earlier research on symptoms prior to SCA has included at least limited patient clinical history (cardiac disease/coronary heart disease and sometimes cardiovascular risk factors),^3–9^ and some analyses have evaluated whether history of heart disease was related to differences in survival,^4^ likelihood of patients’ calling 911 prior to SCA,^5^ or focused on differences in symptom presentation among individuals with coronary artery disease^6^ or hypertrophic cardiomyopathy.^7^ However, to our knowledge, no prior study has evaluated whether the combination of symptoms and specific clinical history was associated with SCA risk.

The literature on prediction of acute coronary events, however, provides proof of concept that a combination of symptoms and clinical phenotype can improve prediction of future events. For example, the HEART score for prediction of acute coronary events for patients presenting with chest pain in the emergency department (ED) included points for risk factors for atherosclerosis, including diabetes, hypertension, hypercholesterolemia, and obesity.^13^ Similarly, a risk score for prediction of acute coronary events developed in Sweden for individuals presenting to the ED with chest pain reported that a combination of symptom characteristics (such as pain triggers, pain location and quality), clinical history (kidney disease, heart failure, atrial fibrillation), labs (Troponin T), and ECG findings differentiated individuals at high vs low risk of acute coronary events.^14^

Our objective in this analysis, therefore, was to evaluate the potential for the combination of patient clinical profile and symptoms to predict ISCA. We employed classification and regression tree (CART) models which are better suited than logistic regression to identify complex, higher-order interactions to potentially detect distinct patient subgroups at the highest risk of ISCA. In the longer term, this data could lead to development of prediction algorithms that could be tested and validated in prospective, general patient populations to answer this question in an urgent care situation: “Is this patient with presenting symptom X and clinical profile Y at a high risk of ISCA?”

## METHODS

### Study Design

We used a case-control design to evaluate whether symptoms combined with patients’ pre-existing clinical comorbidities could distinguish SCA cases from control subjects. All subjects had to have at least one symptom from a pre-determined list. No matching on any other patient characteristics was performed.

### Case and Control Ascertainment and Adjudication

*SCA cases:* Individuals were included from two ongoing community-based studies of out-of-hospital SCA: The Oregon Sudden Unexpected Death Study (SUDS) in the Portland, Oregon metro area (pop ~1 million) and the *Pre*diction of *S*udden Death in Mul*t*i-Ethnic C*o*mmunities (PRESTO) in Ventura County, California (pop ~850,000). All incident cases of presumed SCA were prospectively identified in both studies through collaboration with each county’s 2-tiered EMS system.^15^ Cases underwent detailed adjudication by three staff physicians, based on the EMS pre-hospital care report, medical records from regional hospital systems, death certificates from California or Oregon state vital statistics records, and the Medical Examiner report and autopsy if available. Adjudicated SCA cases were defined as those with a sudden, unexpected pulseless condition of likely cardiac origin.^1^ Cases with an identifiable non-cardiac etiology for cardiac arrest were excluded, such as trauma, drug overdose, and chronic terminal illness (e.g., malignancy not in remission). For this analysis, eligible cases were survivors of SCA aged ≥18 with pre-arrest medical history available and symptom data available from an interview conducted at least one month after the cardiac arrest, or from symptoms recorded by EMS in the pre-hospital care report.

#### Control subjects

The control group was designed to represent individuals ages 18-85 attended by EMS who did not have SCA, but who had symptoms that could have been mistaken for ISCA. Potential control subjects were identified from the list of all 911 calls to each region’s EMS system from Jan 1 through Dec 31, 2019, excluding calls for cardiac arrest, trauma, behavioral calls, and pregnancy complications, and those not resulting in transport to hospital. To select individuals meeting inclusion criteria, we performed a comprehensive automated keyword search of the EMS narrative for any of the pre-defined inclusion symptoms (Supp. Fig. 1) or their synonyms. From this list, we selected successive simple random samples of individuals for manual review of the EMS pre-hospital care narratives and retained those with at least one of the 12 inclusion symptoms and who consented to participate. Control subjects included in this analysis were those with medical history available and who did not have a terminal illness and who participated in an interview.

### Definitions and Source of Symptom Data

Symptom data were obtained from interviews with study subjects and from EMS pre-hospital care records in which the EMS dispatcher and responding paramedics document patient symptoms. Both cases and controls were defined as having a specific symptom if it was reported in either the interview or the EMS narrative. Symptoms of interest were based on a pre-specified list, compiled from symptoms reported in prior studies of SCA^3–9^ and symptoms of acute coronary syndrome^16–18^ (Figure 1). *Interviews:* In the interview, cases were asked, “On the day of your arrest, did you or those around you notice anything wrong [any symptoms] in the minutes or hours before you collapsed?” Control subjects were asked, “On the day of your 9-1-1 call, can you describe what was wrong [any symptoms], in the minutes or hours before your call?” Interviewers entered subjects’ responses according to the list of pre-defined symptoms (Figure 1). *EMS records:* From the EMS pre-hospital care report, trained study staff manually reviewed the narrative text describing the scene of the cardiac arrest (or 911 call for controls) and any symptoms noted by EMS personnel as reported by the patient, family members, or bystanders. For cases, symptoms were required to have clearly preceded the collapse. Signs occurring because of the cardiac arrest (such as gasping / agonal breathing or apparent seizure following collapse) were not recorded as symptoms.

**Figure 1.**
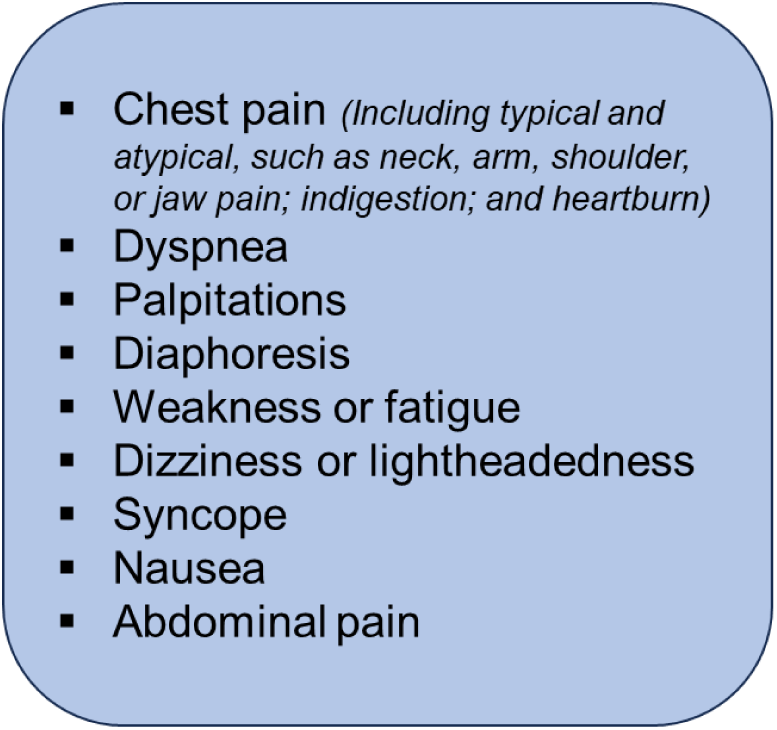
Pre-defined symptoms of interest

**Figure 2.**
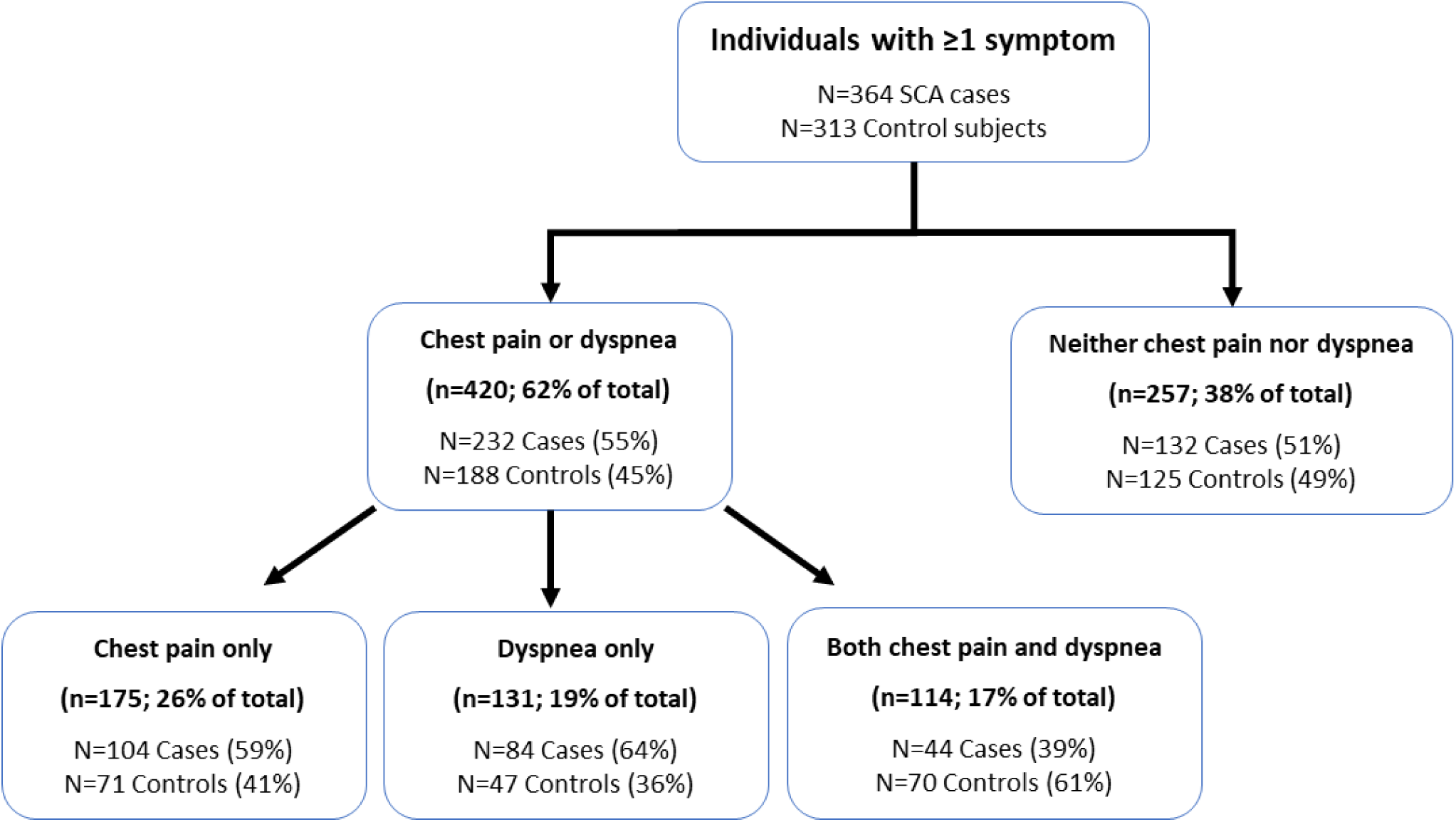
Flow chart of individuals included in analysis. Total subjects included n=677: 364 individuals with sudden cardiac arrest (SCA cases) and 313 individuals without SCA who called 911 and were transported to hospital (Control subjects).

### Source of Medical History Data

Medical history was obtained for all cases and controls by detailed manual review of patients’ existing electronic health records; a clinical condition was considered present if noted by a physician. The PRESTO study was approved by the Institutional Review Boards (IRBs) of Ventura County Medical Center, Cedars-Sinai Health System, and all other participating health systems; the SUDS study was approved by the IRB of Oregon Health and Science University and all other participating health systems. All survivors of cardiac arrest and control subjects provided written informed consent.

### Statistical Analysis

#### Descriptive statistics

We evaluated demographics (age, sex, race/ethnicity), prevalence of existing clinical comorbidities, and individual symptom frequencies (symptoms stratified by sex, based on earlier studies showing sex-specific patterns prior to SCA).^8,12^ Chi-square and *t-*tests with two-sided p-values <0.05 were used to evaluate statistically significant differences using SAS v9.4 (Cary, NC). We then grouped patients into 4 categories based on symptom presentation (chest pain, dyspnea, both, or neither) based on preliminary data and earlier publications indicating chest pain and dyspnea were the two most prevalent symptoms in SCA cases.^12^ Within the four symptom presentation categories, we included seizure as a potential additional predictor of SCA, since it was significant in univariate comparisons and associated with SCA in our earlier publication.^12^

#### Classification and Regression Tree (CART) models

We constructed classification and regression tree (CART) models for the four symptom categories to identify clinical predictors of ISCA among individuals in that symptom group. There were several advantages to using CART models over traditional methods such as logistic regression. CART models are better suited for capturing complex, higher-order interactions. For example, the relationship between heart failure, CAD, chest pain, dyspnea, and SCA risk is unlikely to be purely additive and instead involves subgroup-specific effects that are difficult to explicitly model using conventional regression approaches. Additionally, CART models naturally identify distinct patient subgroups at the highest risk of ISCA, making them particularly useful for risk stratification. Lastly, they offer greater interpretability and clinical applicability compared to regression-based models, as they provide clear decision rules that can be more easily implemented in a clinical setting. We used the R package ‘rpart’ (Rstudio R.4.3.1), setting the complexity parameter at 0.0001 and requiring that each terminal node include at least 10 individuals.^19^ The vector of prior probabilities (defined as the proportion of cases in that symptom category subset) was included in each model to allow for subsequent splits in each CART model to be evaluated against the initial case:control ratio in that symptom category subset, rather than the default of 0.50. We evaluated model performance with the area under the receiver operating characteristic curve (AUC).

#### Predictor variables

Clinical history variables included in CART models included cardiovascular risk factors (diabetes, hypertension (HTN), obesity, hyperlipidemia); established cardiovascular disease (atrial fibrillation, coronary artery disease (CAD), heart failure (HF), peripheral vascular disease (PVD), stroke); and non-cardiac comorbidities (anemia, asthma, cancer, chronic kidney disease, chronic obstructive pulmonary disease (COPD), gastroesophageal reflux disease (GERD), hypothyroid, seizure disorder, sleep apnea, and history of syncope). All CART models also included male sex and seizure symptoms, based on our earlier findings that these symptoms differentiated SCA cases from controls.

#### Model assessment

Following model development, we assessed the stability of each CART model using 5-fold cross-validation. We created 5 random ‘folds’ in each symptom subset through simple random sampling, then fit a CART model in each of 5 training folds (each containing 4/5 of the data, omitting a 1/5 testing fold). Including the same predictor variables as in the full CART models, we assessed the variables selected in each of the 5 training folds. We assessed the AUC in the 4/5 training folds as well as the AUC obtained by applying that model to the 1/5 validation fold. This process allowed (i) evaluation of CART model stability by determining whether similar variables were selected in the 4/5 training datasets compared to the ‘full’ data model, and (ii) evaluation of external prognostic ability using the AUCs of the 5 models in both the 4/5 training and 1/5 validation folds.

## RESULTS

### Participant demographics

Individuals with SCA were more likely to be male than control subjects (71% vs. 47%) (Table 1). SCA cases were also approximately four years older than control subjects and differed somewhat by ethnicity, though this appeared to be largely driven by demographic differences by study site (Supp. Table 1).

**Table 1.** Demographics of individuals with SCA (cases) and a comparison group of individuals (control subjects) with emergency transport to hospital for reasons other than SCA or trauma.

|  | SCA Cases<br>(n=364) | Controls<br>(n=313) |
| --- | --- | --- |
| **Demographics** |  |  |
| Male | 257 (71%) | 146 (47%) |
| Age (mean ± SD), years | 63 ± 14 | 59 ± 15 |
| **Age category** |  |  |
| 18-34 | 7 (2%) | 23 (7%) |
| 35-54 | 93 (26%) | 89 (28%) |
| 55-74 | 187 (51%) | 151 (48%) |
| $\geq 75$ | 77 (21%) | 50 (16%) |
| **Race Ethnicity** |  |  |
| American Indian/Alaska Native | 1 (0.3%) | 1 (0.3%) |
| Asian | 5 (1%) | 9 (3%) |
| Black | 13 (4%) | 20 (6%) |
| Hispanic | 30 (8%) | 58 (19%) |
| Native Hawaiian/Pacific Islander | 3 (1%) | 3 (1%) |
| White Non-Hispanic | 306 (85%) | 221 (71%) |
| Missing | 6 | 1 |

### Patient clinical profile

Both SCA cases and control subjects had a high prevalence of cardiovascular risk factors, including hypertension, diabetes, and obesity (Table 2), though cases had a higher prevalence of hyperlipidemia. Compared to control subjects, SCA cases had a substantially higher prevalence of recognized cardiovascular disease, including atrial fibrillation (30.8% vs. 19.2%, p<0.001), coronary artery disease (52.5% vs. 28.4%), heart failure (40.9% vs. 14.4%), and peripheral vascular disease (12.1% vs. 4.5%) (all p<0.001, Table 2). Stroke history was not significantly different between the two groups (11.0% vs. 9.3%, p=0.46). In contrast, both groups had a similar burden of non-cardiac comorbidities, except that asthma and gastroesophageal reflux disease (GERD) were more common among control subjects (Table 2).

**Table 2.** Medical history, SCA cases and control subjects.

|  | SCA Cases<br>(n=364) | Controls<br>(n=313) | p-value |
| --- | --- | --- | --- |
| **Cardiovascular risk factors** |  |  |  |
| Diabetes | 114 (31.6%) | 98 (31.3%) | 0.94 |
| Hyperlipidemia | 212 (58.2%) | 148 (47.3%) | 0.004 |
| Hypertension | 240 (65.9%) | 207 (66.1%) | 0.96 |
| Obesity | 148 (40.7%) | 114 (36.4%) | 0.26 |
| **Cardiovascular disease** |  |  |  |
| Atrial fibrillation/flutter | 112 (30.8%) | 60 (19.2%) | <0.001 |
| Coronary artery disease | 191 (52.5%) | 89 (28.4%) | <0.001 |
| Heart failure | 149 (40.9%) | 45 (14.4%) | <0.001 |
| Peripheral vascular disease | 44 (12.1%) | 14 (4.5%) | <0.001 |
| Stroke | 40 (11.0%) | 29 (9.3%) | 0.46 |
| **Non-cardiac comorbidities** |  |  |  |
| Anemia | 44 (12.1%) | 50 (16.0%) | 0.14 |
| Asthma | 46 (12.6%) | 69 (22.0%) | 0.001 |
| Cancer | 55 (15.1%) | 63 (20.1%) | 0.09 |
| Chronic kidney disease | 71 (19.5%) | 58 (18.5%) | 0.75 |
| Chronic obstructive pulmonary disease | 61 (16.8%) | 45 (14.4%) | 0.40 |
| Gastroesophageal reflux disease | 66 (18.1%) | 92 (29.4%) | <0.001 |
| Hypothyroid | 48 (13.2%) | 46 (14.7%) | 0.57 |
| Seizure disorder | 11 (3.0%) | 17 (5.4%) | 0.12 |
| Sleep apnea | 54 (14.8%) | 59 (18.9%) | 0.16 |
| Syncope | 39 (10.7%) | 40 (12.8%) | 0.40 |

### Patient symptoms

#### Case-control comparisons among individuals with symptoms

The most common symptoms among male SCA cases were chest pain or discomfort (experienced by 47%) and dyspnea (33%); these symptoms had similar frequencies among male control subjects (44% (p=0.58) and 38% (p=0.32) respectively, Table 3). Seizure-like symptoms (prior to SCA in cases and prior to 911 call in control subjects) were more common among male SCA cases than controls (13% vs. 1%, p<0.001). Among female SCA cases, dyspnea was the most common symptom, experienced by 39%, with a similar frequency (37%) among control subjects (p=0.65, Table 3). Chest pain was less common among female SCA cases (26%) than among female control subjects (46%, p<0.001) (Table 3). As in men, seizure-like symptoms were also more common among female SCA cases than controls (12% vs 5%, p=0.03). All other symptoms (diaphoresis, weakness or fatigue, dizziness or lightheadedness, nausea or vomiting, syncope, palpitations, and abdominal symptoms) were more common among control subjects, with most being statistically significant differences (Table 3).

**Table 3.**
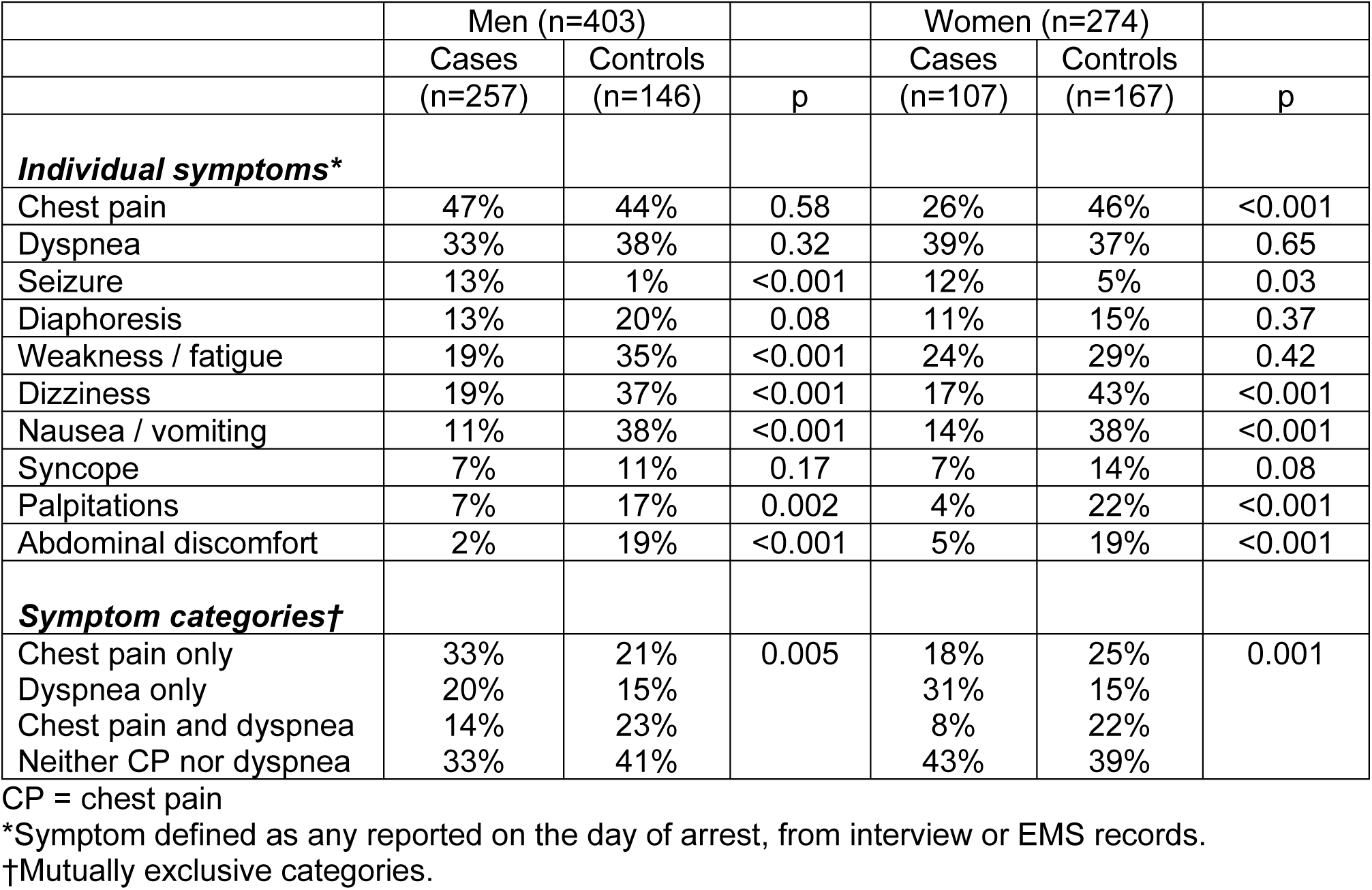
Sex-stratified symptoms among SCA cases and control subjects.

|  | Men (n=403) |  |  | Women (n=274) |  |  |
| --- | --- | --- | --- | --- | --- | --- |
|  | Cases | Controls |  | Cases | Controls |  |
|  | (n=257) | (n=146) | p | (n=107) | (n=167) | p |
| ***Individual symptoms\**** |  |  |  |  |  |  |
| Chest pain | 47% | 44% | 0.58 | 26% | 46% | <0.001 |
| Dyspnea | 33% | 38% | 0.32 | 39% | 37% | 0.65 |
| Seizure | 13% | 1% | <0.001 | 12% | 5% | 0.03 |
| Diaphoresis | 13% | 20% | 0.08 | 11% | 15% | 0.37 |
| Weakness / fatigue | 19% | 35% | <0.001 | 24% | 29% | 0.42 |
| Dizziness | 19% | 37% | <0.001 | 17% | 43% | <0.001 |
| Nausea / vomiting | 11% | 38% | <0.001 | 14% | 38% | <0.001 |
| Syncope | 7% | 11% | 0.17 | 7% | 14% | 0.08 |
| Palpitations | 7% | 17% | 0.002 | 4% | 22% | <0.001 |
| Abdominal discomfort | 2% | 19% | <0.001 | 5% | 19% | <0.001 |
| ***Symptom categories†*** |  |  |  |  |  |  |
| Chest pain only | 33% | 21% | 0.005 | 18% | 25% | 0.001 |
| Dyspnea only | 20% | 15% |  | 31% | 15% |  |
| Chest pain and dyspnea | 14% | 23% |  | 8% | 22% |  |
| Neither CP nor dyspnea | 33% | 41% |  | 43% | 39% |  |

CP = chest pain
\*Symptom defined as any reported on the day of arrest, from interview or EMS records.
†Mutually exclusive categories.

Chest pain was unique in its differential frequency among SCA cases by sex: 47% of male SCA cases vs. 26% of female SCA cases had chest pain. In contrast, male controls and female controls had a similar frequency of chest pain (44% and 46% respectively). Comparing symptoms by sex within study group (male vs. female SCA cases and male vs. female control subjects), all other symptoms were similar by sex, within six percentage points (Table 3).

### CART Models

#### Overview of model results

We created four symptom subsets for CART models: individuals with chest pain only (n=175, 26% of total subjects), with dyspnea only (n=131, 19% of total subjects), with both chest pain and dyspnea (n=114, 17% of total subjects), and with neither chest pain nor dyspnea (n=257, 38% of total subjects) (Figure 1).

The CART models for each symptom subset are shown in Figures 3a-3d (simplified figures) and Supp. Figures 1a-1d (figures of full models). In the four CART models, AUC ranged from 0.728 to 0.813. Sensitivity ranged from 0.59 to 0.85; specificity ranged from 0.55 to 0.81.

**Figure 3.**
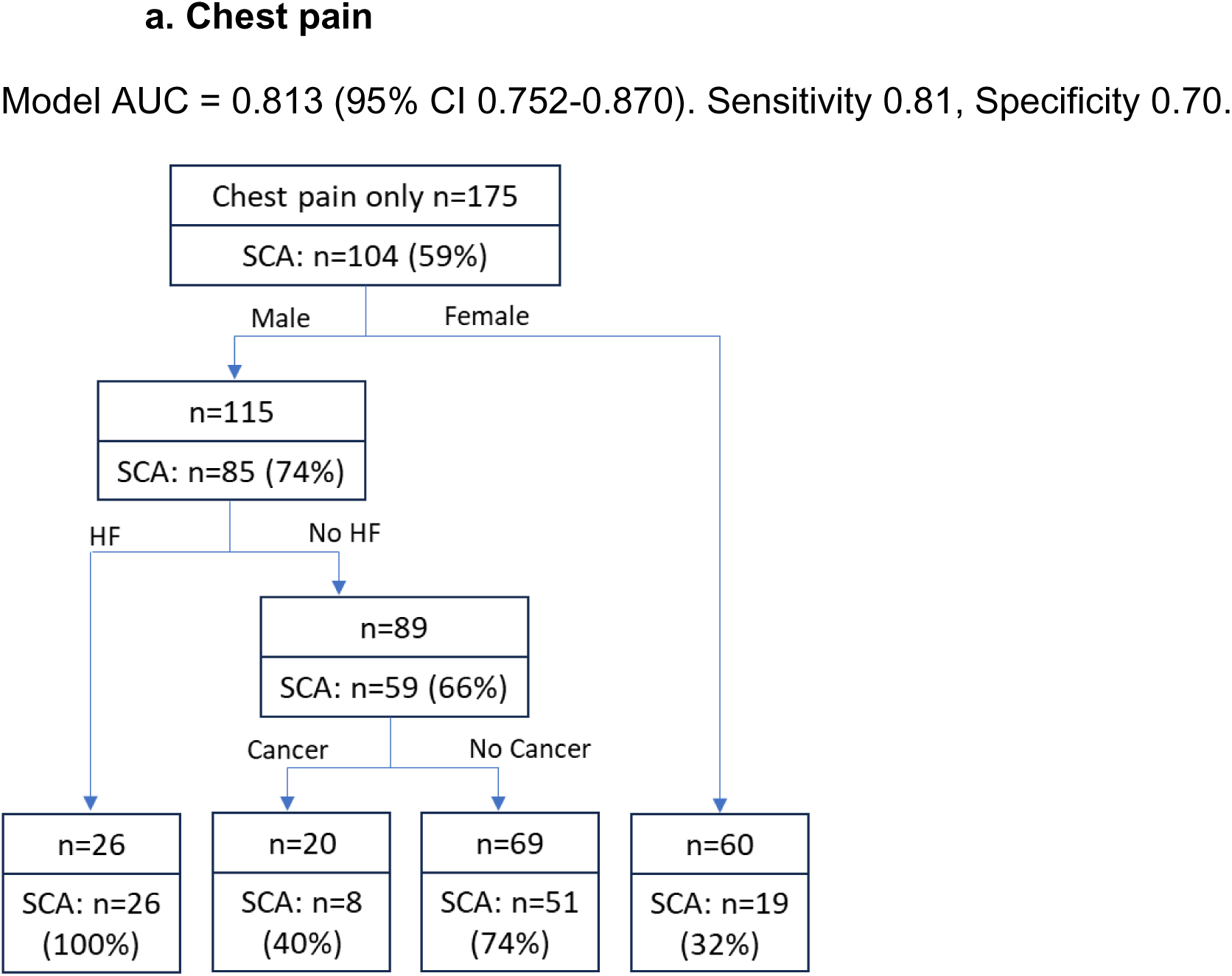
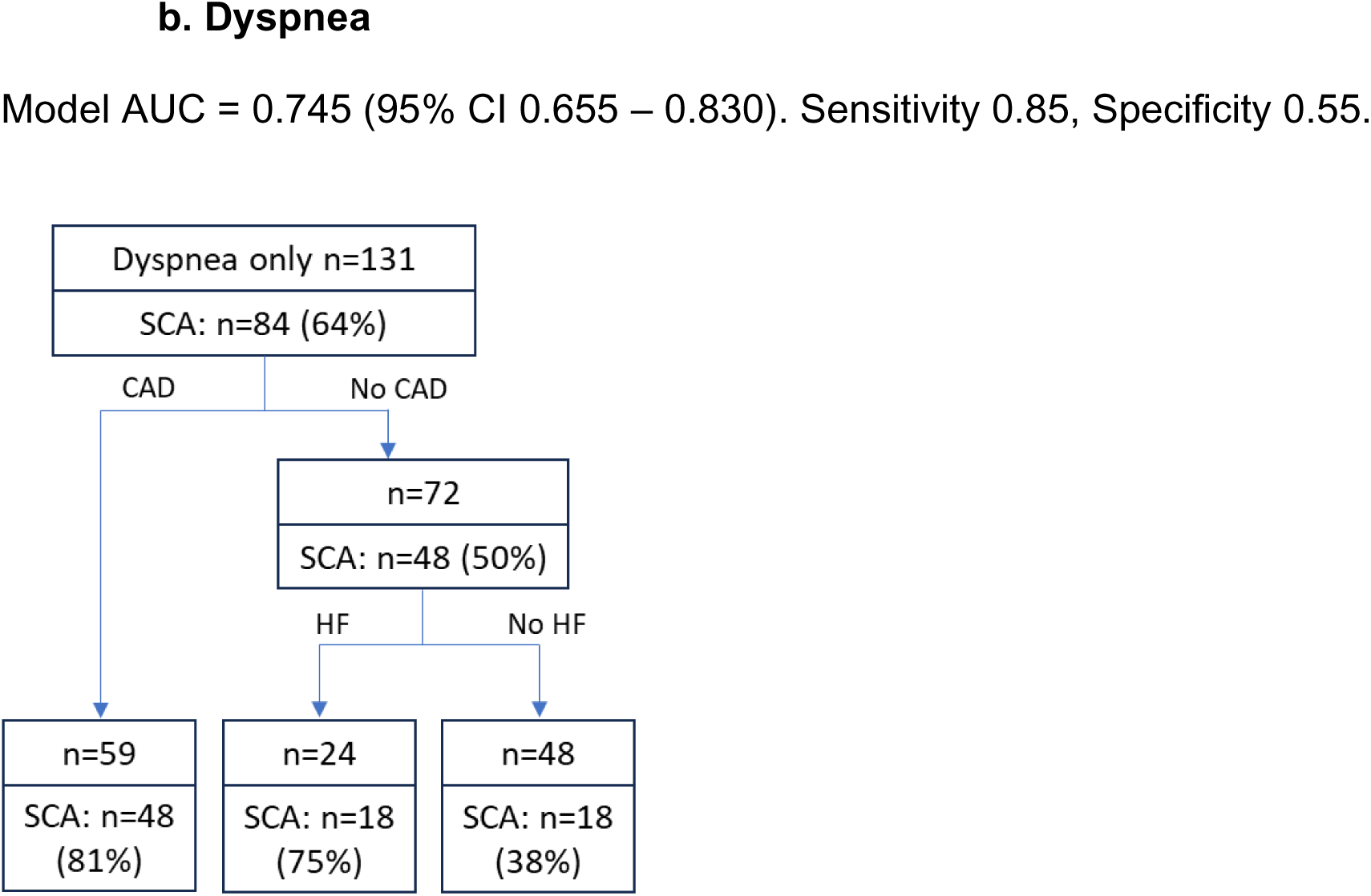
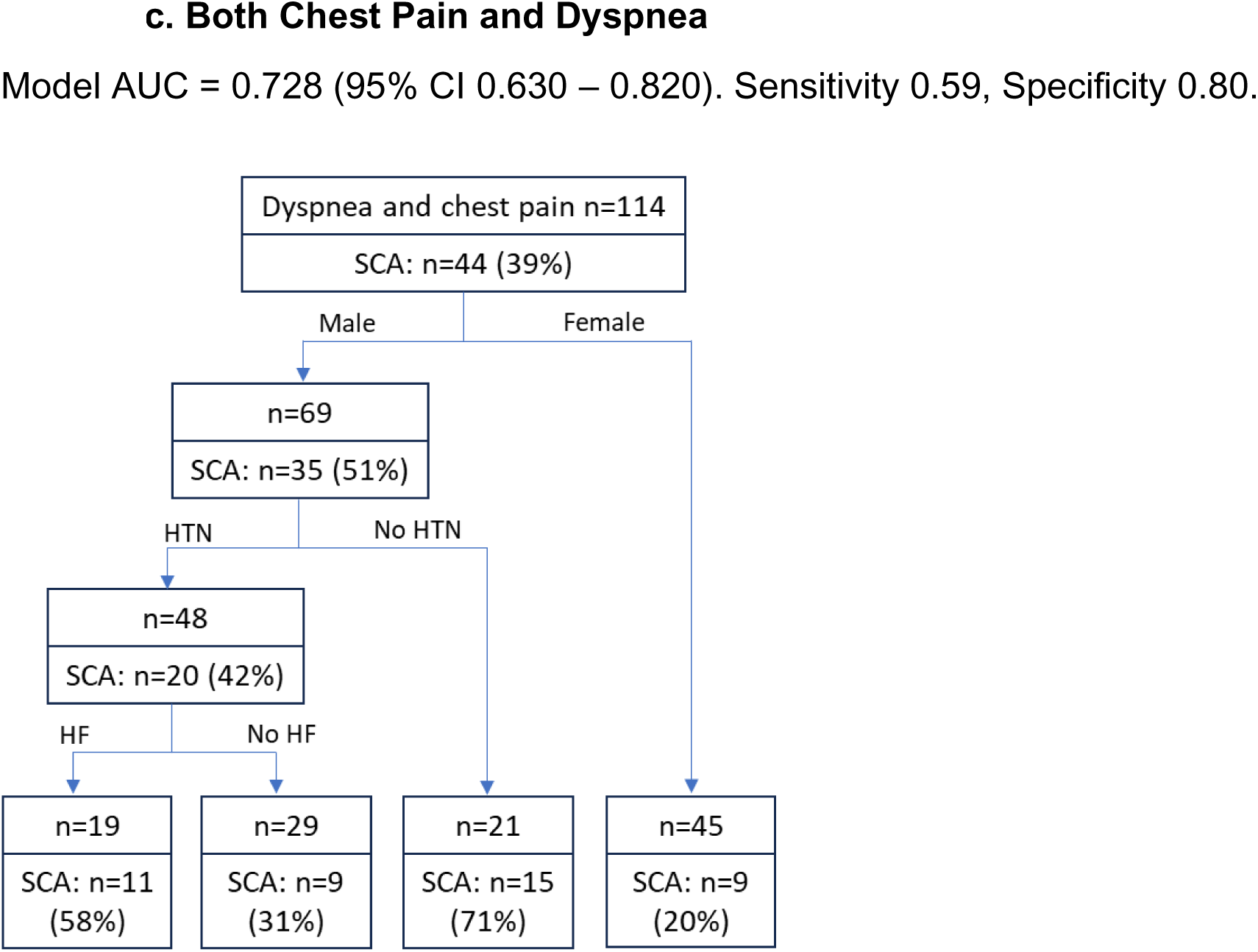
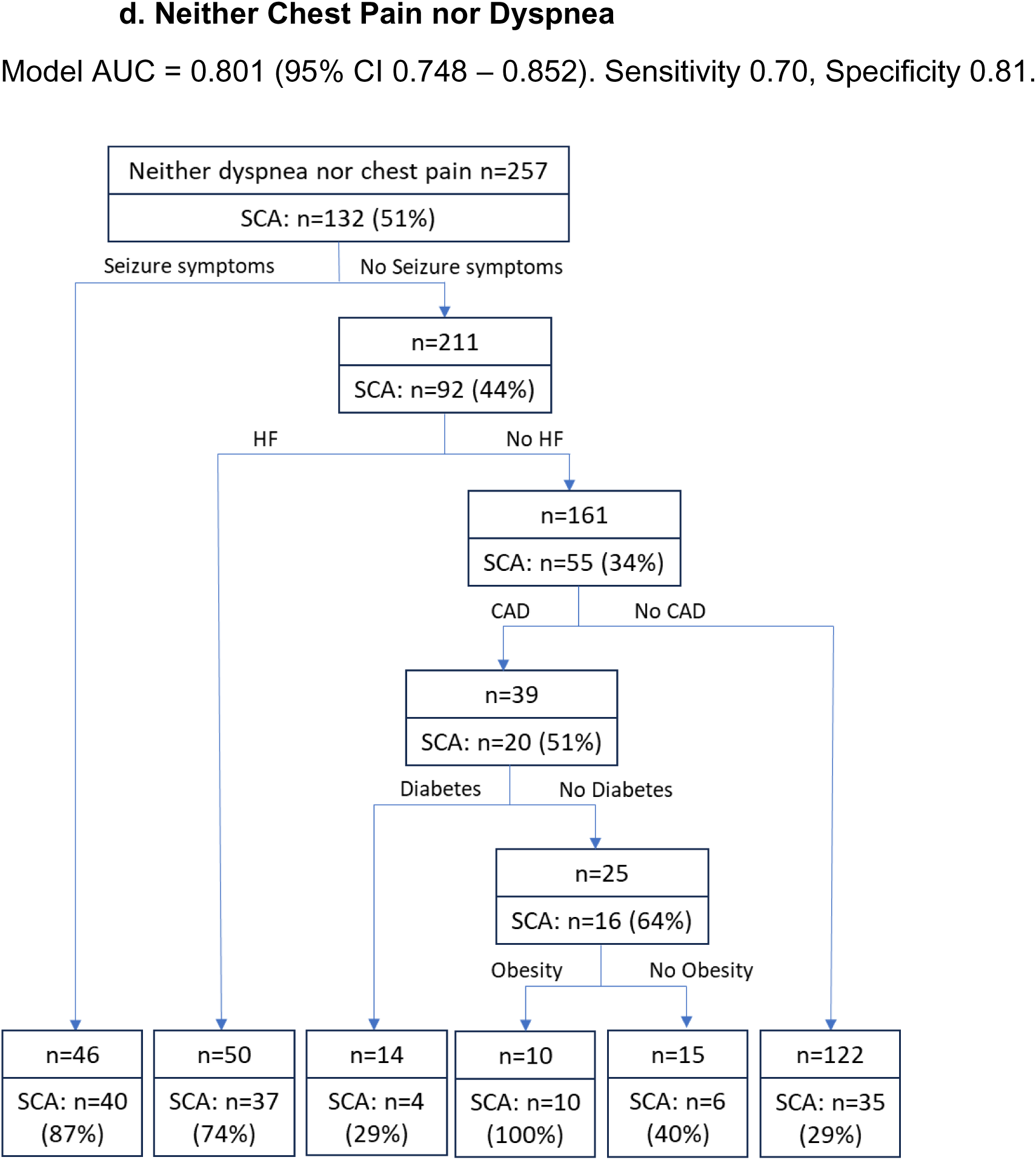
Classification and Regression Trees for prediction of SCA with (a) chest pain, (b) dyspnea, (c) chest pain and dyspnea, (d) neither chest pain nor dyspnea. Number in each top box (node) indicates total subjects (SCA cases and controls). The lower segment of each box indicates the number and proportion of SCA cases among subjects in that node. Features discriminating SCA cases from controls appear in order of importance from top to bottom. Model area under the receiver operating characteristic curve (AUC), sensitivity, and specificity for identification of SCA are noted. *Note: Figures are simplified for clarity by excluding descending branches from a node if each branch included a smaller percentage of SCA cases than the original node*. *Supplemental Figures 1a-1d illustrate the full CART models*.

#### Results by symptom group

In the CART model for chest pain without dyspnea (of whom 59% overall were SCA cases), male sex was the most important classifier. Among men with chest pain, 74% were SCA cases, and among men with HF, 100% were SCA cases (Figure 3a). Among women with chest pain, only 32% were SCA cases (Figure 3a); the likelihood increased only in the small subset of women without cancer but with atrial fibrillation (n=10, 70% of whom were SCA cases) (Supp Figure 1a).

In the model for dyspnea without chest pain (of whom 64% overall were SCA cases), presence of CAD and HF indicated increased likelihood of SCA. Among individuals with a history of CAD, 81% were SCA cases (Figure 3b); among those without CAD but with HF, 75% of individuals were SCA cases. In this symptom group, sex did not differentiate SCA cases from control subjects.

Among individuals presenting with both chest pain and dyspnea, fewer individuals overall (39%) were SCA cases (Figure 3c). Male sex was the most important classifier in this symptom subset (among males, 51% were SCA cases, while among females, 20% were SCA cases). Males with no history of hypertension were more likely to be cases (71%), as were males with both hypertension and HF (58%).

Among individuals with neither chest pain nor dyspnea, 51% overall were SCA cases (Figure 3d). Among individuals who presented with seizure symptoms, 87% were SCA cases. Among those with no seizure symptoms, HF was the most important predictor (74% were SCA cases). Among individuals with no seizure symptoms and no HF, 64% of those with CAD and with no diabetes were SCA cases; this proportion was 100% in the smaller subset of this branch with obesity (Figure 3d).

#### Summary of CART model findings

Table 4 summarizes the CART model findings. Among clinical variables, HF was an important predictor of SCA in every model. Male sex was an important predictor of SCA only when chest pain was present. History of CAD predicted SCA only in the groups without chest pain. Other less important clinical predictors were hypertension, diabetes, absence of cancer history, and obesity. In the group with neither dyspnea nor chest pain, having seizure symptoms the day of arrest was a predictor of SCA risk. For female patients presenting with chest pain (with or without dyspnea), no predictors of SCA emerged.

#### Stability Assessment of CART Models

In 5-fold cross-validation in the 4 symptom subgroups, model average AUCs across the 5 training datasets ranged from 0.748 to 0.785, and across the 5 testing datasets ranged from 0.638 to 0.718. In terms of predictor stability, variables were largely similar but not always in the same order of importance.

## DISCUSSION

In this analysis, a combination of symptoms and clinical profile was able to distinguish individuals with SCA from a comparison group of individuals who did not have SCA. Using CART models to identify patient characteristics associated with higher probability of ISCA, history of HF was an important predictor of SCA among males regardless of symptom presentation and among females only in the absence of chest pain. History of CAD was an important predictor only among individuals without chest pain. Other less important clinical predictors were hypertension, diabetes, absence of cancer history, and obesity. In the group with neither dyspnea nor chest pain, having seizure symptoms the day of arrest predicted SCA risk. For patients presenting with chest pain (with or without dyspnea), male sex was an important predictor of ISCA. For females with chest pain, no predictors of ISCA emerged. These findings could be used to inform development and testing of clinical prediction algorithms for urgent care and emergency settings (EMS, emergency department) in which providers must make critical triage decisions.

To our knowledge, this is the first study to evaluate the combination of presenting symptom(s) with clinical profile to identify individuals who may be at increased risk of ISCA. A substantial number of individuals with SCA have symptoms prior to their arrest,^9^ and there is growing awareness that these symptoms could be useful in preventing ISCA.^10^ While chest pain and dyspnea have consistently been reported as the most common warning symptoms prior to SCA,^4,5,8,9,12^ each of these symptoms, as well as other less common symptoms, are experienced by <40% of individuals, leading to low sensitivity for individual symptoms. The CART models combining symptoms and clinical profile in this analysis achieved AUCs of 0.728 to 0.813, indicating moderately good to good discrimination.

Some studies have characterized the combination of clinical profile and symptoms in patients with acute myocardial infarction (MI) or acute coronary events. Our univariate finding that individuals in our control group had more non-cardiac symptoms (dizziness, weakness, abdominal symptoms, palpitations, and nausea) is consistent with an evaluation of symptom clusters in patients with suspected acute coronary syndrome (ACS) in which patients with a heavy symptom burden or being “weary” were less likely than those with chest symptoms to ultimately receive an ACS diagnosis.^20^ Our findings of less chest pain and higher dyspnea prevalence among women with SCA compared to men with SCA is consistent with a meta-analysis of 27 studies in which women with confirmed ACS had higher odds of dyspnea and lower odds of chest pain.^21^

### Strengths and Limitations

This study’s strengths include the availability of symptom data from both interviews and EMS reports for individuals with SCA and for a comparison group of individuals who also had at least one symptom from a pre-defined set but who did not have SCA. We also had detailed medical history available for all cases and controls. Our use of CART models allowed for identification of patient subgroups at increased risk of SCA based on sex, symptom presentation, and clinical profile. This carefully adjudicated group of SCA cases and unique comparison group allowed evaluation of the question, “Are individuals with presenting symptom X and clinical profile Y likely to have an imminent SCA?”

A few limitations must also be mentioned in evaluating the results of this study. Because this was a case-control study design, it was not possible to calculate a positive or negative predictive value (PPV, NPV) of symptom-comorbidity combinations. The probability of SCA within a decision node could be high, but if the prevalence of the symptom and clinical history combination is low, it could account for a small percentage of total SCA cases and not be particularly useful for clinical prediction. In addition, chest pain and dyspnea are common symptoms, and likely are experienced by many individuals with conditions other than SCA, leading to potentially low specificity. Prospective testing of any symptom and clinical profile combinations is necessary to evaluate its clinical utility in the general population.

Additionally, given the use of a low complexity parameter in CART models, and the multiple subgroup analyses, the results should be interpreted as exploratory and hypothesis generating rather than definitive clinical decision tools. The decision trees highlight potential relationships between symptoms, comorbidities, and SCA risk, but should not be directly applied for risk stratification in practice without further validation. Nonetheless, these models provide a valuable starting point for future research aimed at improving early identification of individuals at imminent risk for SCA.

Other limitations are specific to our patient population. The symptom and clinical profile of survivors of SCA may not be representative of the entire population at high risk for SCA. Finally, defining the appropriate comparison group for SCA is challenging. Our approach was to compare symptomatic individuals with SCA to a group of individuals with at least one of the pre-defined symptoms who also had EMS response and transport but who did not have SCA to determine whether the symptom-clinical profile combination could distinguish them. A different type of control group may have led to different results.

### Conclusions

This analysis demonstrated that a combination of symptoms and clinical profile was able to distinguish individuals with SCA from a comparison group without SCA, with model AUCs ranging from 0.728 to 0.813. History of HF was the most consistent predictor of ISCA; among males HF predicted ISCA regardless of symptom presentation while among females, HF was important only in the absence of chest pain. These models point toward the possibility of identifying individuals at risk of ISCA but need to be evaluated prospectively and potentially augmented with additional features for improved prediction of ISCA.

## Data Availability

De-identified participant data will be made available after publication on reasonable request to the corresponding author, following approval of a proposal and a signed data use agreement.

## Notes

### Competing Interest Statement

The authors have declared no competing interest.

### Funding Statement

Dr. Chugh has received funding from National Institutes of Health, National Heart Lung and Blood Institute Grants R01HL147358 and R01HL145675 for this work. Dr. Chugh holds the Pauline and Harold Price Chair in Cardiac Electrophysiology at Cedars-Sinai, Los Angeles. The funder of the study had no role in study design, data collection, data analysis, data interpretation, or writing of the report.

### Author Declarations

The PRESTO study was approved by the Institutional Review Boards (IRBs) of Ventura County Medical Center, Cedars-Sinai Health System, and all other participating health systems; the SUDS study was approved by the IRB of Oregon Health and Science University and all other participating health systems. All survivors of cardiac arrest and control subjects provided written informed consent.

