## Supplementary Tables 1,2; Supplementary Figures 1(a, b, c, d) for "Prediction of Imminent Sudden Cardiac Arrest Using a Combination of Warning Symptoms and Clinical Features"

**Supplementary Material**

**Supplementary Table 1.** Demographics of SCA cases and control subjects by study site.

| | Oregon SUDS site* | | PRESTO site† | |
|---|---|---|---|---|
| | SCA Cases (n=261) | Controls (n=73) | SCA Cases (n=103) | Controls (n=240) |
| **Demographics** | | | | |
| Male | 185 (71%) | 45 (62%) | 72 (70%) | 101 (42%) |
| Age (mean ± SD), years | 63.4 ± 13.6 | 57.8 ± 13.8 | 62.4 ± 15.3 | 59.9 ± 15.0 |
| **Age category** | | | | |
| 18-34 | 7 (3%) | 6 (8%) | 0 | 17 (7%) |
| 35-54 | 56 (21%) | 20 (27%) | 37 (36%) | 69 (29%) |
| 55-74 | 146 (56%) | 39 (53%) | 41 (40%) | 112 (47%) |
| ≥75 | 52 (20%) | 8 (11%) | 25 (24%) | 42 (18%) |
| **Race Ethnicity** | | | | |
| Am Ind/Alaska Native | 1 (<1%) | 0 | 0 | 1 (<1%) |
| Asian | 1 (<1%) | 1 (1%) | 4 (4%) | 8 (3%) |
| Black/AA | 13 (5%) | 8 (11%) | 0 | 12 (5%) |
| Hispanic | 9 (4%) | 3 (4%) | 21 (20%) | 55 (23%) |
| Native Hawaiian/PI | 2 (1%) | 2 (3%) | 1 (1%) | 1 (<1%) |
| White Non-Hispanic | 229 (90%) | 58 (81%) | 77 (75%) | 163 (68%) |
| Missing | 6 | 1 | 0 | 0 |

*SUDS = Sudden Unexpected Death Study, Portland, OR metro area
†PRESTO = Prediction of Sudden Death in Multi-ethnic Communities, Ventura County, CA

**Supplementary Table 2.** Summary of predictors of SCA in CART models by symptom group.

| Chest Pain only | Chest Pain + Dyspnea | Dyspnea only | Neither Chest Pain nor Dyspnea |
|---|---|---|---|
| Male, HF(+)* | Male, HTN(-) | CAD(+) | Seizure symptom(+) |
| Male, HF(-)* and cancer (-) | Male, HTN(+) and HF(+) | CAD(-) and HF(+) | Seizure symptom(-) and HF(+) |
| | | | Seizure symptom(-), HF(-), CAD(+), diabetes(-), obesity (+) |

*****Plus sign (+) indicates present of condition; minus sign (-) indicates absence of condition.

**Supplementary Figure 1.** Classification and Regression Trees for SCA cases and controls with (a) chest pain, (b) dyspnea, (c) chest pain and dyspnea, (d) neither chest pain nor dyspnea.

Legend: Number in each box (node) indicates total subjects (SCA cases and controls); percentage indicates the proportion of SCA cases among subjects in that node. Nodes shaded green indicates likelihood of SCA higher than in the starting node. Features discriminating SCA cases from controls appear in order of importance from top to bottom of the tree diagram. Model area under the receiver operating characteristic curve (AUC), sensitivity, and specificity for identification of SCA are noted.

*Note: These are the full CART models, including all branches that met CART model specifications.*

**Supp. 1a. Chest pain**

Model AUC = 0.813 (95% CI 0.752-0.870). Sensitivity 0.81, Specificity 0.70.

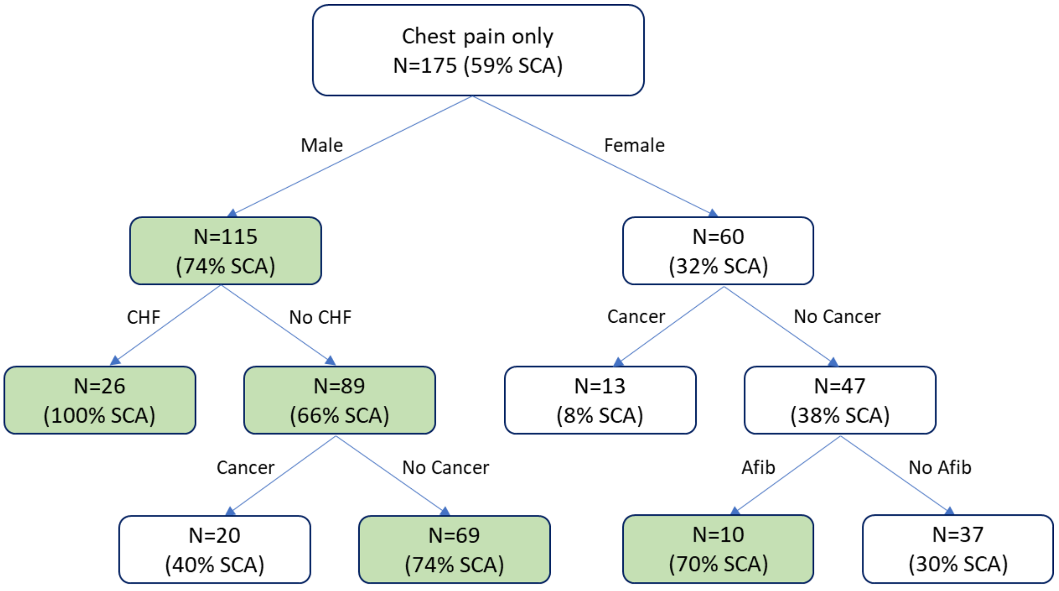

**Supp. 1b. Dyspnea**

Model AUC = 0.745 (95% CI 0.655 – 0.830). Sensitivity 0.85, Specificity 0.55.

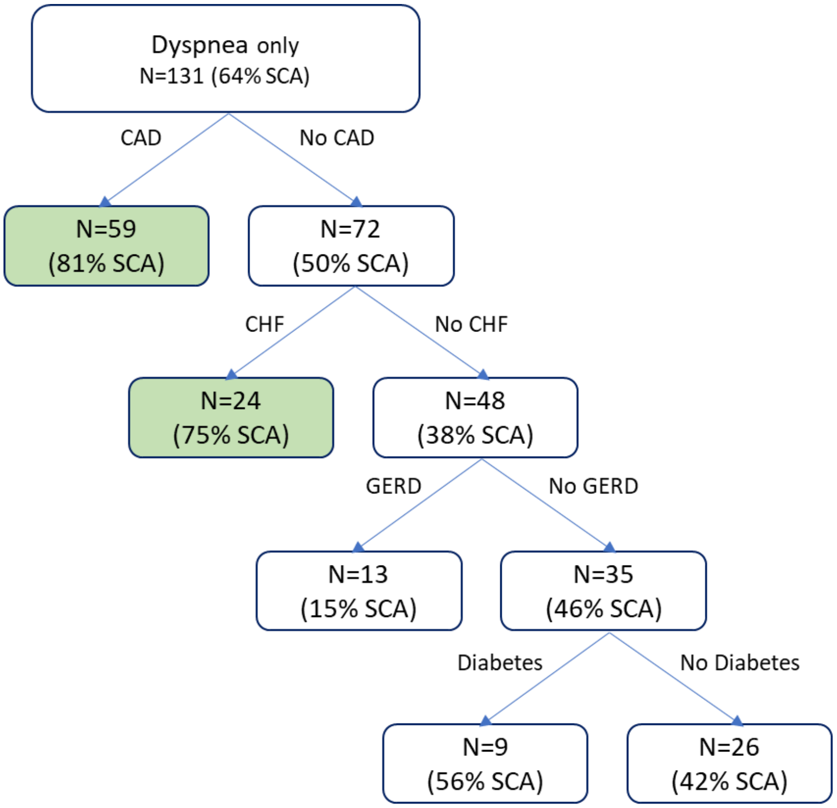

**Supp. 1c. Chest Pain and Dyspnea**
Model AUC = 0.728 (95% CI 0.630 – 0.820). Sensitivity 0.59, Specificity 0.80.

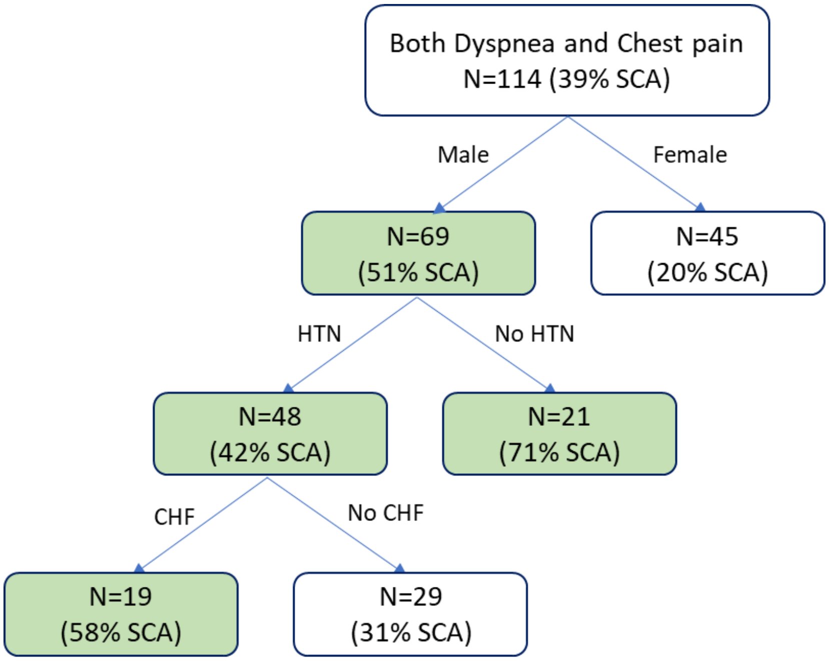

**Supp. 1d. Neither Chest Pain nor Dyspnea**
Model AUC = 0.801 (95% CI 0.748 – 0.852). Sensitivity 0.70, Specificity 0.81.

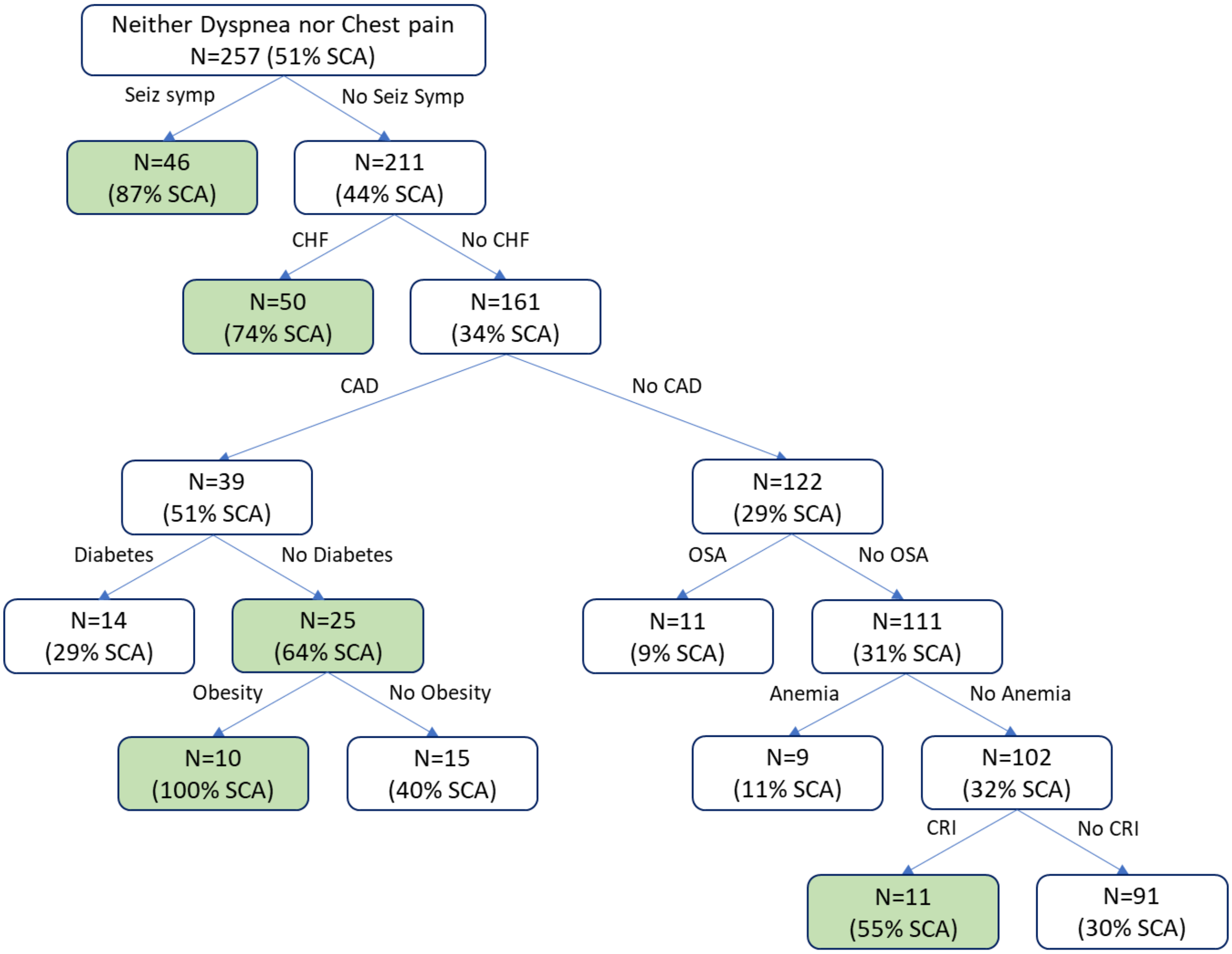
